# Anatomical insights beyond the center edge angle in Borderline Hip Dysplasia: A Computerized tomography study

**DOI:** 10.1101/2024.11.05.24316794

**Authors:** Joaquín Lara, Alejandro Neira, Javier del Río, Alexander Tomic, Alan Garín, Nicolás García, Matías Roby, Carlos De la Fuente

## Abstract

**Background:** Borderline hip dysplasia (BhD) might be associated with insufficient acetabular coverage. Thus, we investigated potential differences in acetabular anatomical measurements derived from computerized tomography (CT) that characterize BhD compared with healthy controls.

**Methods:** BhD patients (LCEA between 18º and 25º) and healthy controls (LCEA between 25º and 40º) underwent anteroposterior pelvic X-ray and CT to study Wiberg and Tönnis angle, extrusion and Fear index, notch width and depth, anterior and posterior wall height, anterior and posterior articular surfaces, articular circumference, the ratio between the anterior articular surface and the articular circumference, the ratio between the posterior articular surface and the articular circumference, and the ratio between the notch width and the articular circumference. An independent two-tailed t-test, U of Mann-Whitney test, and odds ratios were obtained (α = 5%).

**Results:** Twenty-three BhD patients aged 31.5 ± 8.3 years and LCEA 21.6° ± 4.0° and thirty-one healthy controls aged 34.1 ± 8.0 years and LCEA 33.7° ± 5.5° were included. The sensitive CT features for BhD were anterior (*p* < 0.001) acetabular surface, anterior (*p* = 0.009), anteroposterior (*p* = 0.008) and posterior (*p* < 0.008) acetabular surface ratios, and acetabular notch width (*p* = 0.002).

**Conclusions:** BhD CT characterization provides tridimensional anatomical insights beyond the LCEA. BhD insufficient acetabular coverage can be found along the superior (lower Wiberg angle, and increased Tönnis angle and extrusion index), anterior (lower anterior acetabular surface and anteroposterior acetabular surface ratio, and increased posterior acetabular ratio), and inferior (increased acetabular notch width) axis.

## 1 Introduction

Hip dysplasia is an underlying tridimensional bone architecture problem that increases stress concentration (Vaudreuil and McClincy, 2020; Freiman *et al*., 2022), causing labral tearing, cartilage attrition (Freiman *et al*., 2022), and early end-stage osteoarthritis (Brooks, 2013; Cheng *et al*., 2023). Clinically is characterized by hip pain and functional limitations (Cooperman, Wallensten and Stulberg, 1983; Cheng *et al*., 2023). The lateral center edge angle (LCEA) or Wiberg angle (Vaudreuil and McClincy, 2020; Dornacher *et al*., 2023) is the standard radiology to measure the lack of lateral femoral coverage as the main common finding in severe hip dysplasia (Cheng *et al*., 2023; Dornacher *et al*., 2023). The coverage quantification using anteroposterior pelvis radiography allows the hip dysplasia clsssification between (LCEA < 17º) and normal coverage (LCEA > 25°) (Vaudreuil and McClincy, 2020). Nevertheless, 18º to 25º of LCEA represents a lateral Borderline acetabular coverage (Tönnis and Heinecke, 1999; Dornacher *et al*., 2023) that mischaracterizes the original tridimensional coverage architecture, overshadowing other morphology alterations (McClincy *et al*., 2019; Dornacher *et al*., 2023). Unfortunately, Borderline hip dysplasia (BhD) can affect 19.8% to 23.3% of the asymptomatic general population and 12.8% of the symptomatic patients (Freiman *et al*., 2022).

Although the LCEA has served as a primary parameter for evaluating and guiding surgical interventions in severe hip dysplasia (Wells *et al*., 2017), the LCEA alone does not provide a comprehensive understanding of pathological hip changes (Cheng *et al*., 2023) because the LCEA only represents one hip morphology aspect. This structural underrepresentation may favor inconsistent and variable treatment choices (Cheng *et al*., 2023) and compromise patient outcomes.

On the other hand, orthopedic literature has described diverse BhD patterns (Dornacher *et al*., 2023). For example, lateral under coverage, normal lateral coverage, and lateral and posterior under coverage have previously been described as common patterns (Graesser *et al*., 2020; Dornacher *et al*., 2023). However, the anterior coverage deficit is another frequent finding (McClincy *et al*., 2019; Zimmerer *et al*., 2020; Dornacher *et al*., 2023), which has been associated with worse joint restoration after periacetabular osteotomy (Kitamura *et al*., 2022). Further to BhD patterns, a past report suggested that BhD would be well characterized by anterior femoral coverage morphology deficits (Jacobsen, Rømer and Søballe, 2006). However, this is still under discussion (Beaulé, 2019), and extended morphological descriptors related to anteroposterior coverage like notch width and depth, anterior and posterior wall height, anterior and posterior articular surfaces, articular circumference, the ratio between the anterior articular surface and the articular circumference, the ratio between the posterior articular surface and the articular circumference, and the ratio between the notch width and the articular circumference would show intrinsical anatomical characteristics of BhD patients. Consequently, computerized tomography (CT) might help to explore the acetabular BhD architecture beyond the LCEA (Jessel *et al*., 2009; Siebenrock *et al*., 2013).

Due to the scarce morphological three-dimensional BhD characterization, we aim to investigate potential differences in acetabular anatomical measurements derived from CT that characterize BhD compared with healthy controls. Secondly, we determine the odds ratios for the CT anatomical features with statistical significance. We hypothesize that notch width and depth, anterior and posterior wall height, anterior and posterior articular surfaces, articular circumference, the ratio between the anterior articular surface and the articular circumference, the ratio between the posterior articular surface and the articular circumference, and the ratio between the notch width and the articular circumference characterize distinctive patterns in BhD compared to healthy controls.

## 2 Material and Methods

### 2.1. Study design and setting

In this case-control and cross-sectional study, we prospectively recruited participants from the same institution between 2018 and 2023, according to sample size estimations. From patients between 18 and 40 years, we included two independent groups recruited in parallel. The BhD and healthy group were defined for an LCEA between 18º and 25º, and 25º to 40º, respectively. All participants were studied with anteroposterior pelvic X-ray and CT. Notch width and depth, anterior and posterior wall height, anterior and posterior articular surfaces, articular circumference, the ratio between the anterior articular surface and the articular circumference, the ratio between the posterior articular surface and the articular circumference, and the ratio between the notch width and the articular circumference were compared between groups.

Finally, this study followed the declaration of Helsinki regarding the ethical principles of Human experimentation, was approved by the governmental province ethical committee [*Blinded for revision*], written informed consent was obtained from all participants, and this study followed the STROBE statement to report this original research.

### 2.2. Participants

This study included patients with clinical records of BhD treated by the same orthopedic team, aged between 18 and 40 years, with the capacity to walk, upstairs and downstairs without assistance, a positive test for anterior hip pain and abnormal foot progression angle walking, and anteroposterior pelvic X-ray records. Healthy participants were included if they did not have a dysplasia diagnosis, were between 18 and 40 years old, and had the capacity to walk upstairs and downstairs without assistance. The exclusion criteria for both groups were any history of the spine, pelvis and lower limb fractures, chronic musculoskeletal disorders, soft tissue alterations, and any cardiorespiratory and neurologic pathology. The exclusion criteria for patients were any orthopedic conditions different from dysplasia, leg length difference > 1.5 cm, and no habitual use of orthopedic shoes to correct the length difference during walking. The exclusion criteria for the healthy group were a positive orthopedic test for anterior hip pain, no normal foot progression angle walking, and no radiological sign of osteoarthritis.

*A priori* estimated sample size was made for a two-tailed independent t-test for two groups, a large effect size (0.857) obtained from mean values of the anterior uncovered (0.25) and covered (0.46) index, from the estimated mean dispersion (0.245) previously described (Dornacher *et al*., 2023), an alpha of 5%, and a statistical power set at 80%. The minimum total estimated sample was 46 participants or 23 participants by each group, obtaining a critical t-value of 2.02. The estimation was made using the G*Power software 3.1.9.2 (Universität Kiel, Germany).

### 2.3. Procedures

One of four orthopedic surgeons specialists in hip preservation clinically evaluated patients and previous records and images of dysplasia also were checked. At this time point, the sociodemographic data were collected (age, weight, and sex), and new anteroposterior pelvis X-ray and CT images were requested (LCEA, Wiberg and Tönnis angle, and Extrusion and fear index were obtained). From whom obtained an LCEA < 25º and fulfilling all inclusion and exclusion criteria, orthopaedic surgeons confirmed the dysplasia diagnosis. Healthies participants were invited to participate from the community and only if they had records in our clinic. Patients and healthies participants underwent a CT study using a 16-slice CT (Brivo CT 385 series, G.E. Healthcare, U.S.A) with a 0.62 mm resolution and pixel spacing of 0.43 mm × 0.43 mm for research purposes.

Manually (mean value of three measurements), a blinded senior musculoskeletal radiologist and orthopedic surgeon measured ten specific acetabulum anatomy features derived from axial CT scans, previously described. The measurements of CT images were realized in the horizontal plane through the center of the femoral head. Center of the femoral head was determined with visual exploration using horizontal-coronal linked and synchronized views to ensure the highest femoral diameter. All images data were exported into DICOM format and analyzed in Philips Radiology Information System software (version 11,Philips MEdical Systems, Netherlands).

### 2.4. Outcomes

The anatomical measurements are summarized in Figure 1 and defined as follow: 1) acetabular notch width defined by the distance between the posterior border of anterior lunate surface and the anterior border of posterior lunate surface measured in millimeters, 2) acetabular fossa depth defined as the distance drawn from the midpoint of the acetabular notch width line with a perpendicular line to the bottom of the notch measured in millimeters, 3) Anterior lunate wall height defined as the distance from the anterior edge of acetabular fossa to the posterior border of anterior lunate surface measured in millimeters, 4) Posterior lunate wall height defined as the distance from the posterior edge of acetabular fossa to the anterior border of posterior lunate surface measured in millimeters, 5) Anterior lunate surface width defined as the distance between anterior and posterior borders of anterior lunate surface measured in millimeters, 6) Posterior lunate surface defined as the distance between anterior and posterior borders of posterior lunate surface measured in millimeters, 7) Total lunate surface defined as the sum of anterior and posterior lunate surfaces, 8) Anterioposterior lunate surface ratio (AP ratio) defined as the ratio of the anterior and posterior lunate surfaces, 9) Lunate anterior/total surface ratio (AT ratio) defined as the ratio of the anterior and total lunate surfaces, 10) Lunate posterior/total surface ratio (AP ratio) defined as the ratio of the anterior and total lunate surfaces.

**Figure 1.**
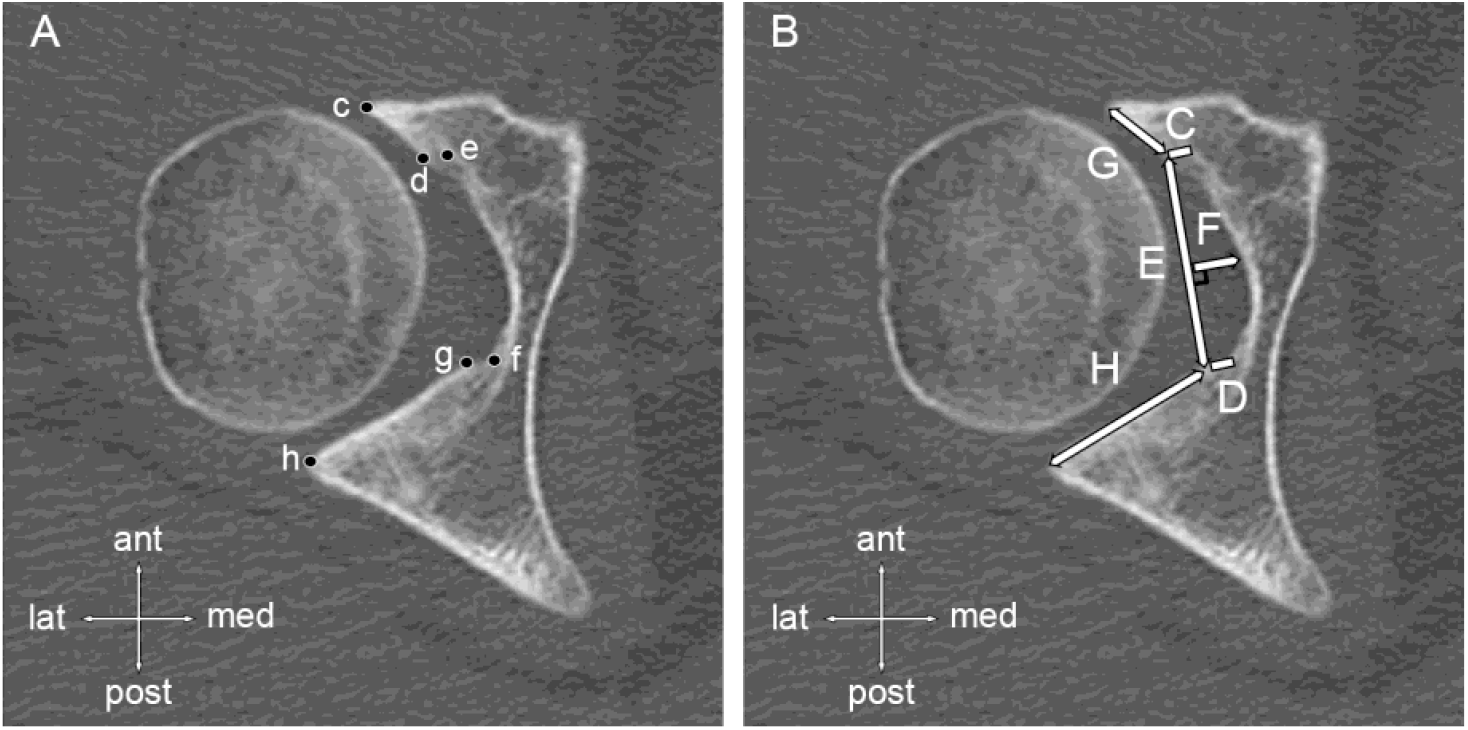
CT variables landmarks and measurements in reference axial hip plane. A. CT landmarks are shown in c as the anterior border of the anterior lunate surface, d as the posterior border of the anterior lunate surface, e as the anterior edge of the acetabular fossa, f as the posterior edge of the acetabular fossa, g as the anterior border of posterior lunate surface, and has the posterior border of the posterior lunate surface. B. CT distances measured in this study are shown in C as anterior lunate wall heigth, D as posterior lunate wall heigth, E acetabular notch width, F acetabular fossa depth, G as anterior lunate surface, and H as posterior lunate surface.

### 2.4. Statistical Analysis

Normality and homoscedasticity principles were checked through Shapiro-Wilk and Levene’s tests. Consequently, data were described as the mean and standard deviation for variables distributed normally with homogeneity. Variables not distributed normally or with homoscedasticity were described as median and interquartile range.

The comparison between groups was made using an independent two-tailed t-test for variables distributed normally with homogeneity, while a U of Mann-Whitney test was made for variables not distributed normally or with no homogeneity. Effect sizes (Hedges’ g for different sample sizes) and confidence intervals were described. We used the standardized interpretation as small for 0.2 and 0.5, moderate for 0.5 and 0.8, and large for values higher than 0.8.

The odds ratios were determined for acetabular anatomical features with statistical significance with respect to the Wiberg angle. The effect size was estimated as the ln(odds)/1.81 (Chinn, 2000), confidence intervals for odds ratios, and *p*-values of the one-tailed Fisher exact test were also estimated. All significance level was set at α = 5%. All statistical analyses were performed using SPSS 20.0 software (I.B.M. Inc., U.S.A.).

We calculated the intraclass correlation coefficient (ICC) and the 95% confidence interval to determine the intra- and inter-observer CT measurements reliability. Two separate time points, at least two weeks apart with blinded clinical information, were performed in an independent sample of 10 participants (α = 5% and β= 20%) (Bujang, 2017).

## 3 Results

Fifty-four participants were included in this study. Twenty-three patients and thirty-one participants were included in the BhD and healthy control groups, respectively. There were eight dropouts in the BhD group. Three men and twenty women aged 31.5 ± 8.3 years and LCEA 21.6° ± 4.0° characterized the BhD group. Ten men and twenty-one women aged 34.1 ± 8.0 years and LCEA 33.7° ± 5.5° characterized the healthy control group. Radiological measurements of groups are described in Table 1.

**Table 1.** Radiological and computerized tomography (CT) outputs of the study.

| Radiological measurements | Borderline Dysplasia | Healthy Control | $\Delta$ | 95%CI | Effect Size |
| --- | --- | --- | --- | --- | --- |
| Wiberg Angle,<sup>o</sup> | 23.0 [3]<sup>z</sup> | 34.0 [6] | 12.1 | 9.5 – 14.7 | 2.5 |
| Tönnis Angle,<sup>o</sup> | 12.0 ± 4.4<sup>z</sup> | 7.8 ± 2.7 | 4.2 | 2.3 – 6.1 | 1.2 |
| Extrusion Index, % | 68.0 [13]<sup>z</sup> | 83.0 [8] | 19.5 | 12.3 – 26.7 | 1.7 |
| Fear Index,<sup>o</sup> | 6 [6] | 10.0 [8] | 0.7 | -5.0 – 6.3 | 0.1 |
| CT measurements | Borderline Dysplasia | Healthy Control | $\Delta$ | 95%CI | Effect Size |
| Acetabular Notch Width, mm | 31.0 [6.1]<sup>Ψ</sup> | 27.7 [3] | 3.2 | 1.0 – 5.1 | 0.9 |
| Acetabular Fossa Depth, mm | 9.3 [2.6] | 8.3 [1.5] | 0.4 | -1.0 – 0.4 | 0.3 |
| Anterior Lunate wall height, mm | 3.4 ± 0.7 | 3.4 ± 1.3 | 0.03 | -0.51 – 0.62 | 0.03* |
| Posterior Lunate wall height, mm | 4.4 [2.3] | 4.2 [1.4] | 0.8 | -0.3 – 2.0 | 0.4 |
| Anterior Lunate surface, mm | 12.1 ± 2.1<sup>z</sup> | 15.0 ± 3.0 | 2.9 | 1.4 – 4.4 | 1.1 |
| Posterior Lunate surface, mm | 21.8 ± 3.3 | 23.5 ± 3.6 | 1.8 | -0.2 – 3.7 | 0.5 |
| AP Lunate surface ratio, mm/mm | 0.56 ± 0.1<sup>Ψ</sup> | 0.64 ± 0.1 | 0.08 | 0.02 – 0.14 | 0.8 |
| Lunate Anterior/Total surface ratio, mm/mm | 0.35 ± 0.04<sup>Ψ</sup> | 0.39 ± 0.05 | 0.03 | 0.01 – 0.06 | 0.8 |
| Lunate Posterior/Total surface ratio, mm/mm | 0.64 ± 0.04<sup>Ψ</sup> | 0.61 ± 0.05 | 0.02 | 0.01 – 0.06 | 0.8 |
| Total Lunate surface, mm | 39.1 ± 4.2 | 38.5 ± 5.7 | 1.4 | -3.3 – 2.1 | 0.1 |

Data are described as mean ± standard deviation and Median [interquartil range].
<sup>Ψ</sup>: Statistical significance for Displasia Group compared with Controls ( $p < 0.05$ ).
<sup>z</sup>: Statistical significance for Displasia Group compared with Controls ( $p < 0.001$ ).
\*: two decimal was shown to appreciate the magnitude of the effect size.
95%CI: 95% of confident interval for mean differences between groups.
$\Delta$ : Mean difference between groups.
CT: computerized tomography.
AP: antero-posterior.
<sup>o</sup>: sexagesimal degree.

The patients’ acetabular CT images are shown in Figure 2. There was a lower Wiberg angle (*p* < 0.001; large effect size), Anterior (*p* < 0.001; large effect size) acetabular surface, Anterior (*p* = 0.009; large effect size), and Anteroposterior (*p* = 0.008; large effect size) acetabular surface ratio in the BhD group compared with healthy controls (Table 1). In contrast, there was a higher Tönnis angle (*p* < 0.001; large effect size), extrusion index (*p* < 0.001; large effect size), acetabular notch width (*p* = 0.002; large effect size), and posterior acetabular surface ratio (*p* < 0.008; large effect size) in the BhD group compared with healthy controls (Table 1).

**Figure 2.**
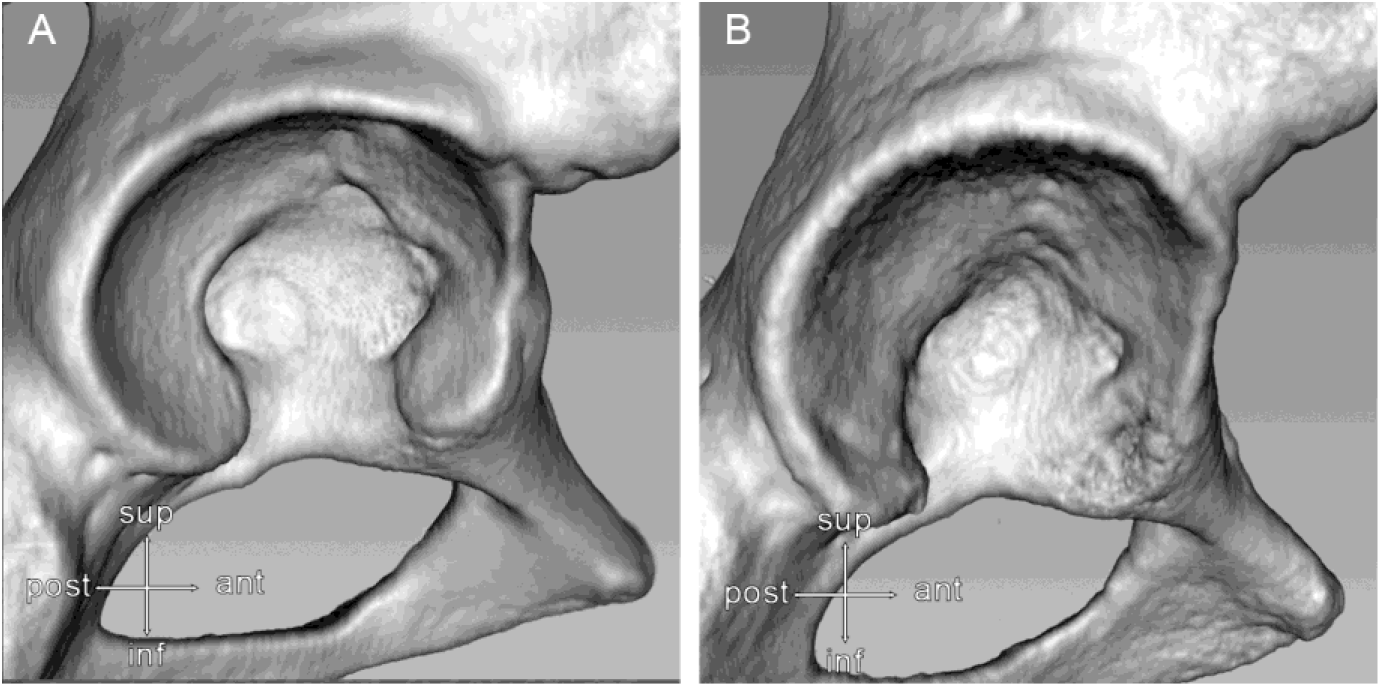
Three-dimensional CT image renderization of the Acetabulum. The images illustrates the anatomical differences observed in the anterior lunate surface and acetabular notch width with a preserved posterior lunate surface between a healthy subject (A) and a BhD patient (B).

The odd ratio for acetabular notch width was 2.1 [95%IC: 0.69 – 6.66] with *p* = 0.147 and effect size of 0.41 (small effect size), for anterior acetabular surface was 11.2 [95%IC: 2.23 – 56.17] with *p* = 0.001 and effect size of 1.33 (large effect size), for anteroposterior acetabular surface ratio was 2.8 [95%IC: 0.83 – 8.54] with *p* = 0.083 and effect size of 0.57 (moderate effect size), for anterior acetabular surface ratio was 3.0 [95%IC: 0.94 – 9.71] with *p* = 0.053 and effect size of 0.61 (moderate effect size), and for posterior acetabular surface ratio was 3.0 [95%IC: 0.94 – 9.71] with *p* = 0.053 and effect size of 0.61 (moderate effect size). The intra- and inter-observer reliability measurements are described in Table 2.

**Table 2.** Reliability measures of the outcomes.

|  | Intra-observer |  | Interobserver |  |
| --- | --- | --- | --- | --- |
|  | Borderline<br>Dysplasia | Healthy<br>Control | Borderline<br>Dysplasia | Healthy<br>Control |
|  | ICC (95% CI) | ICC (95% CI) | ICC (95% CI) | ICC (95% CI) |
| **CT measurements** |  |  |  |  |
| Acetabular Notch Width, *mm* | 0.36 (0.70-0.81) | 0.98 (0.93-0.99) | 0.53 (0.36-0.87) | 0.71 (0.01-0.92) |
| Acetabular Fossa Depth, *mm* | 0.83 (0.34-0.95) | 0.93 (0.75-0.98) | 0.78 (0.22-0.94) | 0.80 (0.24-0.95) |
| Anterior Lunate wall height, *mm* | 0.83 (0.26-0.95) | 0.92 (0.71-0.98) | 0.77 (0.17-0.94) | 0.89 (0.39-0.97) |
| Posterior Lunate wall height, *mm* | 0.82 (0.35-0.95) | 0.96 (0.86-0.99) | 0.79 (0.24-0.94) | 0.93 (0.74-0.98) |
| Anterior Lunate surface, *mm* | 0.93 (0.73-0.98) | 0.98 (0.92-0.99) | 0.83 (0.38-0.95) | 0.92 (0.73-0.98) |
| Posterior Lunatesurface, *mm* | 0.94 (0.77-0.98) | 0.98 (0.91-0.99) | 0.89 (0.56-0.74) | 0.96 (0.87-0.99) |
| AP Anterioposterior Lunate surface ratio, *mm/mm* | 0.96 (0.86-0.92) | 0.98 (0.94-0.99) | 0.91 (0.62-0.98) | 0.94 (0.78-0.98) |
| Lunate Anterior/Total surface ratio, *mm/mm* | 0.95 (0.80-0.98) | 0.98 (0.94-0.99) | 0.88 (0.48-0.97) | 0.94 (0.78-0.98) |
| Lunate Posterior/Total surface ratio, *mm/mm* | 0.95 (0.80-0.98) | 0.98 (0.93-0.96) | 0.88 (0.48-0.97) | 0.95 (0.88-0.98) |
| Total Lunate surface, *mm* | 0.93 (0.72-0.98) | 0.99 (0.96-0.99) | 0.84 (0.4-0.96) | 0.96 (0.87-0.99) |

*AP: antero-posterior.*
*ICC: Intraclass correlation coefficient.*
*CI: Confident interval.*
*mm: millimeter.*
*%: percent.*

## 3 Discussion

The main finding of our study was that BhD patients develop anatomical pathological characteristics related to diminished: 1) anterior acetabular coverage (lower anterior acetabular surface and anteroposterior acetabular surface ratio, and increased posterior acetabular ratio), 2) lateral acetabular coverage (lower Wiberg angle, and increased Tönnis angle and extrusion index), and 3) inferior acetabular coverage (increased acetabular notch width), and 4) our study highlights there is between 11.2 to 2.8 and 2.1 times for BhD patients have anterior and inferior acetabular coverage insufficiency, respectively. These identified anatomical insufficiencies would compromise the anterior, superior, and inferior joint stability given by insufficient structural (rigid) constraints, promoting a pathological acetabular stress distribution(Vaudreuil and McClincy, 2020) and non-physiological femoral head migration into the joint, similar to micro-instability or severe osteoarthritis previously described (Resnick, 1975; Hayward *et al*., 2014; Eijer and Hogervorst, 2017; Tazawa *et al*., 2019). Based on our findings, acetabular reorientation techniques like periacetabular osteotomy importantly should primarily aim for a three-dimensional reorientation and coverage augmentation without over-coverage to normalize the joint stability and the stress distribution (Clohisy *et al*., 2008) to prolong the preservation of the Acetabulum, instead primarily non-structural repair and reconstructions tissues (secondary stabilizers). Here, we showed a novelty finding of a lack of inferior acetabular coverage due to an insufficient anterior acetabular wall that was not previously observed.

The lower anterior acetabular surface and anteroposterior acetabular surface ratio, and increased posterior acetabular ratio were sensitive to show diminished anterior acetabular coverage in our study. Our finding of insufficient anterior acetabular coverage for BhD patients is in accordance with anatomical findings in female dysplasia (Nepple *et al*., 2017; McClincy *et al*., 2019). But here, we were able to determine that the anterior acetabular surface was the most sensitive variable with the highest effect size and odd ratio. This last statement expresses that the ratio of having or not having diminished anterior acetabular coverage is 11.2 times that of BhD patients compared to healthy controls. In this sense, BhD patients have a high risk (21/23 [91.3%] patients) of developing insufficient anterior acetabular coverage.

The acetabular notch width was found to be altered more significantly in BhD patients than in healthy controls. Our findings suggest that insufficient anterior acetabular coverage influences the appropriate inferior coverage, increasing the notch width. Compared to healthy controls, the ratio of developing or not developing an increased acetabular notch width is 2.1 times for BhD patients compared to healthy controls. In this sense, BhD patients also have a risk (16/23 [69.6%] patients) of developing insufficient inferior acetabular coverage. The acetabular notch and the transverse acetabular ligament are physiological barriers against inferior dislocation (Nomani *et al*., 2023). The transverse acetabular ligament with the labrum partially supports the inferior load-bearing for the femoral head movements (Beverland, 2010). Therefore, an increased acetabular notch width would be an important new source of micro-instability or hypermobility (Resnick, 1975; Hayward *et al*., 2014; Eijer and Hogervorst, 2017; Tazawa *et al*., 2019), which, as far as we know, has not been discussed previously for BhD patients. Surgical planning addressing inferior hip stability as a probable anatomical alteration in BhD patients is relevant and should be considered for better tridimensional reconstruction and appropriate coverage.

This study highlights that BhD patients have an increased risk of developing both superior, anterior, and inferior acetabular coverage insufficiency based on the obtained odds ratios. An improved and personalized anatomical patient study better might orientate the tridimensional reconstruction surgery to restore hip stability. Nowadays, biomechanical simulations for stress distribution restoration, tridimensional visualization, printing, and artificial intelligence support for patient classification would help surgeons in the pre-surgical planning and management of BhD patients. Another future effort is to find algorithms that allow the projection of sensitive CT features to traditional dysplasia radiology. Finally, adequate anatomical structure restoration of the Acetabulum should be the primary aim before advancing with secondarily joint stabilizers.

We know our study is not out of limitations. The main limitation here is that our study mainly included female BhD patients. Thus, the pattern associated with males (posterolateral lack of coverage) was not found in the present study. The CT scanning reduces the weight-bearing capacity, which is reproduced by X-ray images. Finally, cadaveric and biomechanical simulations need to explore the intensity and consequence of instability associated with BhD patients in future setups.

## 4 Conclusion

BhD CT characterization provides tridimensional anatomical insights beyond the LCEA. The CT identified acetabular anatomical features sensitive for BhD patients were the anterior acetabular surface, the acetabular notch width, and the anterior, anteroposterior and posterior acetabular surface ratios. BhD insufficient acetabular coverage can be found along the superior (lower Wiberg angle, and increased Tönnis angle and extrusion index), anterior (lower anterior acetabular surface and anteroposterior acetabular surface ratio, and increased posterior acetabular ratio), and inferior (increased acetabular notch width) axis.

## Data Availability

All data produced in the present study are available upon reasonable request to the authors.

## References

Beaulé, P.E. (2019) Editorial Commentary: Quantifying Anterior and Lateral Acetabular Coverage in Hip Dysplasia: What About Posterior Coverage?, Arthroscopy: The Journal of Arthroscopic & Related Surgery: Official Publication of the Arthroscopy Association of North America and the International Arthroscopy Association, 35(4), pp. 1117–1119. Available at: 10.1016/j.arthro.2019.01.041.

Beverland, D. (2010) The transverse acetabular ligament: optimizing version, Orthopedics, 33(9), p. 631. Available at: 10.3928/01477447-20100722-22.

Brooks, P.J. (2013) Dislocation following total hip replacement: causes and cures, The Bone & Joint Journal, 95-B(11 Suppl A), pp. 67–69. Available at: 10.1302/0301-620X.95B11.32645.

Bujang, M.A. (2017) A simplified guide to determination of sample size requirements for estimating the value of intraclass correlation coefficient: a review | Semantic Scholar, Archives of Orofacial Sciences, 12, pp. 1–11.

Cheng, H. et al. (2023) Can we determine anterior hip coverage from pelvic anteroposterior radiographs? A study of patients with hip dysplasia, BMC musculoskeletal disorders, 24(1), p. 522. Available at: 10.1186/s12891-023-06624-2.

Chinn, S. (2000) A simple method for converting an odds ratio to effect size for use in meta-analysis, Statistics in Medicine, 19(22), pp. 3127–3131. Available at: 10.1002/1097-0258(20001130)19:22<3127::aid-sim784>3.0.co;2-m.

Clohisy, J.C. et al. (2008) A systematic approach to the plain radiographic evaluation of the young adult hip, The Journal of Bone and Joint Surgery. American Volume, 90 Suppl 4(Suppl 4), pp. 47–66. Available at: 10.2106/JBJS.H.00756.

Cooperman, D.R., Wallensten, R. and Stulberg, S.D. (1983) Acetabular dysplasia in the adult, Clinical Orthopaedics and Related Research, (175), pp. 79–85.

Dornacher, D. et al. (2023) Acetabular deficiency in borderline hip dysplasia is underestimated by lateral center edge angle alone, Archives of Orthopaedic and Trauma Surgery, 143(7), pp. 3937–3944. Available at: 10.1007/s00402-022-04652-6.

Eijer, H. and Hogervorst, T. (2017) Femoroacetabular impingement causes osteoarthritis of the hip by migration and micro-instability of the femoral head, Medical Hypotheses, 104, pp. 93–96. Available at: 10.1016/j.mehy.2017.05.035.

Freiman, S.M. et al. (2022) Prevalence of Borderline Acetabular Dysplasia in Symptomatic and Asymptomatic Populations: A Systematic Review and Meta-analysis, Orthopaedic Journal of Sports Medicine, 10(2), p. 23259671211040455. Available at: 10.1177/23259671211040455.

Graesser, E. et al. (2020) DEFINING THE BORDERLINE HIP: HIGH VARIABILITY IN ACETABULAR COVERAGE AND FEMORAL DEFORMITY, Orthopaedic Journal of Sports Medicine, 8(4 suppl3), p. 2325967120S00210. Available at: 10.1177/2325967120S00210.

Hayward, M.F. et al. (2014) Reliability of the Interprofessional Collaborator Assessment Rubric (ICAR) in Multi Source Feedback (MSF) with post-graduate medical residents, BMC Medical Education, 14(1), p. 1049. Available at: 10.1186/s12909-014-0279-9.

Jacobsen, S., Rømer, L. and Søballe, K. (2006) The other hip in unilateral hip dysplasia, Clinical Orthopaedics and Related Research, 446, pp. 239–246. Available at: 10.1097/01.blo.0000201151.91206.50.

Jessel, R.H. et al. (2009) Radiographic and patient factors associated with pre-radiographic osteoarthritis in hip dysplasia, The Journal of Bone and Joint Surgery. American Volume, 91(5), pp. 1120–1129. Available at: 10.2106/JBJS.G.00144.

Kitamura, K. et al. (2022) Effect of coronal plane acetabular correction on joint contact pressure in Periacetabular osteotomy: a finite-element analysis, BMC Musculoskeletal Disorders, 23, p. 48. Available at: 10.1186/s12891-022-05005-5.

McClincy, M.P. et al. (2019) Mild or Borderline Hip Dysplasia: Are We Characterizing Hips With a Lateral Center-Edge Angle Between 18° and 25° Appropriately?, The American Journal of Sports Medicine, 47(1), pp. 112–122. Available at: 10.1177/0363546518810731.

Nepple, J.J. et al. (2017) Three Patterns of Acetabular Deficiency Are Common in Young Adult Patients With Acetabular Dysplasia, Clinical Orthopaedics and Related Research, 475(4), pp. 1037–1044. Available at: 10.1007/s11999-016-5150-3.

Nomani, K. et al. (2023) Complete Ossification of Transverse Acetabular Ligament - Embryological and Clinical Perspective, La Clinica Terapeutica, 174(4), pp. 326–330. Available at: 10.7417/CT.2023.2445.

Resnick, D. (1975) Patterns of migration of the femoral head in osteoarthritis of the hip. Roentgenographic-pathologic correlation and comparison with rheumatoid arthritis, The American Journal of Roentgenology, Radium Therapy, and Nuclear Medicine, 124(1), pp. 62–74. Available at: 10.2214/ajr.124.1.62.

Siebenrock, K.A. et al. (2013) Valgus hip with high antetorsion causes pain through posterior extraarticular FAI, Clinical Orthopaedics and Related Research, 471(12), pp. 3774–3780. Available at: 10.1007/s11999-013-2895-9.

Tazawa, M. et al. (2019) Superior migration of the femoral head in patients with severe hip osteoarthritis influences the gait patterns of the coronal plane, Hip International: The Journal of Clinical and Experimental Research on Hip Pathology and Therapy, 29(4), pp. 446–451. Available at: 10.1177/1120700019827250.

Tönnis, D. and Heinecke, A. (1999) Acetabular and femoral anteversion: relationship with osteoarthritis of the hip, The Journal of Bone and Joint Surgery. American Volume, 81(12), pp. 1747–1770. Available at: 10.2106/00004623-199912000-00014.

Vaudreuil, N.J. and McClincy, M.P. (2020) Evaluation and Treatment of Borderline Dysplasia: Moving Beyond the Lateral Center Edge Angle, Current Reviews in Musculoskeletal Medicine, 13(1), pp. 28–37. Available at: 10.1007/s12178-020-09599-y.

Wells, J. et al. (2017) Femoral Morphology in the Dysplastic Hip: Three-dimensional Characterizations With CT, Clinical Orthopaedics and Related Research®, 475(4), pp. 1045–1054. Available at: 10.1007/s11999-016-5119-2.

Zimmerer, A. et al. (2020) Is Hip Arthroscopy an Adequate Therapy for the Borderline Dysplastic Hip? Correlation Between Radiologic Findings and Clinical Outcomes, Orthopaedic Journal of Sports Medicine, 8(5), p. 2325967120920851. Available at: 10.1177/2325967120920851.

